# Awareness and preference for HPV self-collection among Aboriginal and Torres Strait Islander women and people with a cervix

**DOI:** 10.64898/2025.12.25.25343011

**Authors:** Louise E Mitchell, Emily Phillips, Chloe J Jennett, Claire Bavor, Tessa Saunders, Claire Nightingale, Megan A Smith, Lisa J Whop, Screen Your Way Investigator Team

**Affiliations:** Yardhura Walani, National Centre for Aboriginal and Torres Strait Islander Wellbeing Research, Australian National University, Canberra, ACT, Australia; Sydney School of Public Health, The University of Sydney, Sydney, NSW, Australia; Evaluation and Implementation Science Unit, Centre for Health Policy, Melbourne School of Population and Global Health, University of Melbourne, Melbourne, VIC, Australia

## Abstract

This study evaluated awareness and preference for Human Papillomavirus (HPV) self-collection for cervical screening among Aboriginal and Torres Strait Islander screen-eligible participants. A whole-of-population online survey was conducted between December 2023 and April 2024, with responses from 555 Aboriginal and Torres Strait Islander women and people with a cervix. Around 80% of survey participants had heard of self-collection, with healthcare providers the most mentioned source of this knowledge. Never-screened participants were less likely to have heard of self-collection and more likely to report traditional (n=18, 23%) and social media (n=12, 15%) as sources of knowledge of self-collection compared to their healthcare provider (n=13, 16%). Most never-screened participants said self-collection would make them more likely to have a cervical screening test (n=59, 75%). Overall, 275 participants indicated a clear preference for self-collection (50%). Screening history was significantly associated with a preference for self-collection, with Aboriginal and Torres Strait Islander women and people with a cervix who did not screen regularly (adjusted odds ratio [adjOR]=1.9, 95% confidence interval [95%CI]=1.2–3.0, p<0.01), who had never screened (adjOR=3.5, 95%CI=1.8–6.9, p<0.001), or who lived in regional or remote areas (ajdOR=1.6, 95%CI=1.0–2.5, p=0.03) significantly more likely to prefer self-collection. Age, educational attainment and sexuality did not influence preference in the model. These findings indicate a clear preference for self-collection among under- and never-screened Aboriginal and Torres Strait Islander women and people with a cervix. Despite this, never-screened participants were less likely to have heard of this option. There is an opportunity to reach clients who have never screened or are overdue for cervical screening by raising awareness of self-collection. Other than remoteness, sociodemographic characteristics were not associated with a preference for self-collection, reinforcing the need to offer all eligible clients the choice of collection methods.

## Introduction

Australia is on track to become the first to actively eliminate cervical cancer (1, 2), however, there are inequities relating to socio-economic status, remoteness of residence and by Indigenous status. (3) To reach the elimination threshold, cervical cancer incidence rates for Aboriginal and Torres Strait Islander women and people with a cervix must be reduced by 67%. (4) This requires a significant increase in screening among under- and never-screened individuals and targeted efforts to overcome program inequities to ensure elimination includes Aboriginal or Torres Strait Islander women and people with a cervix. (5)

In December 2017, Australia’s National Cervical Screening Program (NCSP) transitioned from two-yearly cytology-based tests to five-yearly Human Papillomavirus (HPV) DNA-based primary screening which enabled the option of self-collected vaginal samples. Initially, concerns about the sensitivity of self-collected samples (6) meant it was offered under a restricted model leading to barriers such as low clinician uptake and poor awareness among eligible participants. (7, 8) This resulted in an estimated fewer than 1% of those eligible using self-collection in the initial two years, with uptake among Aboriginal and Torres Strait Islander people unknown. (9) Following strong evidence showing that self-collected samples are as sensitive as clinician-collected ones and modelling that indicated increased participation would yield greater benefits than any potential loss in sensitivity, Australia revised its policy in July 2022 to offer self-collection to all eligible individuals - referred to as ‘universal access’. (10–12). This policy change aimed to reduce barriers to participation and address inequities in cervical cancer outcomes (13), as self-collection offers participants greater control, privacy, comfort, and convenience. (14)

While recent participation data are not available for Aboriginal and Torres Strait Islander women and people with a cervix, research indicates that self-collection is highly acceptable to Aboriginal and Torres Strait Islander peoples, supports more flexible, culturally responsive delivery models to improve access and therefore has the potential to increase participation in cervical screening. (14–17) We have previously reported on survey findings of the experience of Aboriginal and Torres Strait Islander women and people with a cervix who had recently screened, since the policy change offering universal access to self-collection.(REFERENCE – Under Review - PONE-D-25-29330) Among 261 recently screened Aboriginal and Torres Strait Islander women and people with a cervix, 58% were offered a choice of collection method, and two-thirds of those screened using self-collection. For those who collected their own sample, the main reasons were it was less embarrassing, they felt more in control of their body, and it was less scary. However, many were not offered a choice, and only half felt adequately informed, highlighting the need to consistently provide options and support informed decision-making. We have not previously reported on the experiences of Aboriginal and Torres Strait Islander women and people with a cervix who do not screen, including whether they are aware of self-collection or would prefer this option, nor on the characteristics of those most likely to prefer self-collection.

This study aimed to quantify awareness of HPV self-collection and identify factors associated with a preference for self-collection among screen-eligible Aboriginal and Torres Strait Islander women and people with a cervix, including those who do not currently screen.

## Materials and Methods

Methods and materials have been previously described in (REFERENCE – Under Review - PONE-D-25-29330). We outline the previously described methods and present the data analysis approach for this component of the study.

### Positionality

This work is led by a research team with Aboriginal and Torres Strait Islander lived experience (LJW, EP), Aboriginal and Torres Strait Islander leadership in public health (LJW, EP), epidemiology (LM, LJW, MAS), experience working in Aboriginal and Torres Strait Islander health (LM, LJW, EP) and extensive experience in cervical cancer prevention (all authors).

### Governance

Cultural governance is monitored by Aboriginal and Torres Strait Islander Reference Group, Thiitu Tharrmay, at the Australian National University. Thiitu Tharrmay in Ngiyampaa language translates as ’to share/exchange knowledge’. An Aboriginal and Torres Strait Islander Project Caucus comprised of Aboriginal and Torres Strait Islander Chief and Associate Investigators provided over-arching project governance to ensure that the project is culturally safe and meets the needs and priorities of Aboriginal and Torres Strait Islander people and communities.

### Ethics approval

Ethics approval was obtained from the Australian Institute of Aboriginal and Torres Strait Islander Studies Research Ethics Committee (REC-0209) and ACON (RERC 202404) and was ratified by The Australian National University (H/2023/1434) and the University of Melbourne (2023-28454-47328-1) Human Research Ethics Committees. Individual consent was obtained from all study participants. Before beginning the survey, participants were shown an embedded consent form within Qualtrics. Written informed consent was obtained by asking participants the question: “Would you like to take part in this survey?” Only those who selected “I agree to take part in this survey” were able to proceed, thereby indicating their consent to participate.

### Survey development

An electronic survey was designed containing questions about awareness and experience of, and future preferences for, self-collection cervical screening. The survey was designed to capture sociodemographic variables (including Indigenous status, age, sex, gender, sexuality, highest level of education, and postcode) and comprehensive feedback on cervical screening practices and preferences. For respondents who had recently screened (i.e. since July 2022), questions asked about their experience at their most recent screen, including if they were offered the choice of self-collection, how they chose to screen, and were they given sufficient information to support their choice. Questions were primarily multiple-choice questions to ensure ease of response and consistency in data collection; however, free text fields were included to capture any specific reasons or additional comments. Questions were informed by the COM-B system for behaviour change (18) and Theoretical Domains Framework. (19) The participant information and consent forms and survey were pilot tested by both non-Indigenous and Indigenous peoples, including a non-Indigenous consumer advisory group. The survey was hosted on Qualtrics and took approximately 10-15 minutes to complete.

### Recruitment

Participants were recruited to the study through multiple channels, including a paid Meta campaign via the Cancer Council Australia Instagram and Facebook page, targeted promotion through peak Aboriginal and Torres Strait Islander Community Organisations, and through local stakeholder groups, social media, websites, newsletters, and hard-copy flyers. Recruitment was open between 14 December 2023 and 13 April 2024. Participants accessed the survey through an anonymous link and received a plain language statement. Participants were not renumerated for their time but at the completion of the survey, participants were invited to enter a draw to win a voucher valued at AUD$100. To maintain confidentiality, one study team member (CJ) managed a separate database for the voucher draw, ensuring that survey responses remained independent from participant contact details; 12 vouchers were available exclusively for Aboriginal and Torres Strait Islander participants. Random selection function was used in Excel for the prize draw. The prize draw was completed within 1 month from the final dataset being downloaded, and participant contact details were deleted following this.

### Participants

In total, 11,311 people consented to the survey, of which 677 identified as Aboriginal and/or Torres Strait Islander. Of these, 2 people were aged <24 or >74, 19 whose sex was not recorded female at birth, and 21 who answered no/unsure to having a cervix were ineligible and therefore excluded. A review of data quality, informed by published studies (20, 21) and data available to the investigators from the Qualtrics platform, for identifying potentially fraudulent responses was undertaken. We excluded surveys less than 40% complete (i.e. did not reach key questions on outcome variables) or completed in <120 seconds. We then undertook manual review of surveys utilising Qualtrics fraud detection data; ReCaptca score of <0.5 and Fraud ID >30, and groups of >5 survey which were submitted within a 1-minute window, which identified 191 responses. The manual process involved review of key free text fields (where did you hear about this survey [asked twice], qualitative response questions, and email address), some of which were specifically included to help identify fraudulent responses. Two co-authors undertook the manual process, and 80 responses were excluded on consensus (EP and CJ). This analysis reports on the 555 Aboriginal and Torres Strait Islander people who were eligible for the study. Results for non-Indigenous participants will be reported separately.

## Data Analysis

The analytical approach was grounded in Rigney’s Indigenist research methodology (22) under the guidance of established Aboriginal and Torres Strait Islander governance processes through Yardhura Walani. These principles ensured that Aboriginal and Torres Strait Islander peoples led and informed decisions about data collection, interpretation, and use. Data ownership remains with participants, with the research team acting as custodians.

Research questions included: were participants aware of the option of self-collection? Did participants prefer self-collection? If so, what participant level factors were associated with this preference?

Postcode was used to classify the participant’s place of residence by geographic remoteness using Australian Statistical Geography Standard Remoteness Structure derived from Accessibility/Remoteness Index of Australia Plus (ARIA+) (23), and area-level relative disadvantage using Socio-Economic Indexes for Areas (SEIFA) Index of Relative Socioeconomic Disadvantage 2021 (24).

Participants were categorised as being regular screeners, not-regular screeners, or never screened based on reported screening history, frequency, age and sexual history as per Fig 1. Participants were classified as having a clear preference for self-collection (either collecting the sample themselves or assisted by a healthcare provider using a self-collection swab without a speculum) as per Fig 2. Survey question design limited the ability to distinguish preferences beyond self-collection among never-screened participants; thus, ’clinician collection’, ’either’, and ’unsure’ responses (Q4.9 and Q4.10) were grouped to create a binary option of Other/Unsure. While an alternative model comparing self-collection to clinician-collection was considered, it would exclude never-screened participants—who could only select self-collection – and remove screening history from the model. Given the likely influence of screening history on self-collection preference, this approach was not adopted.

**Fig 1.**
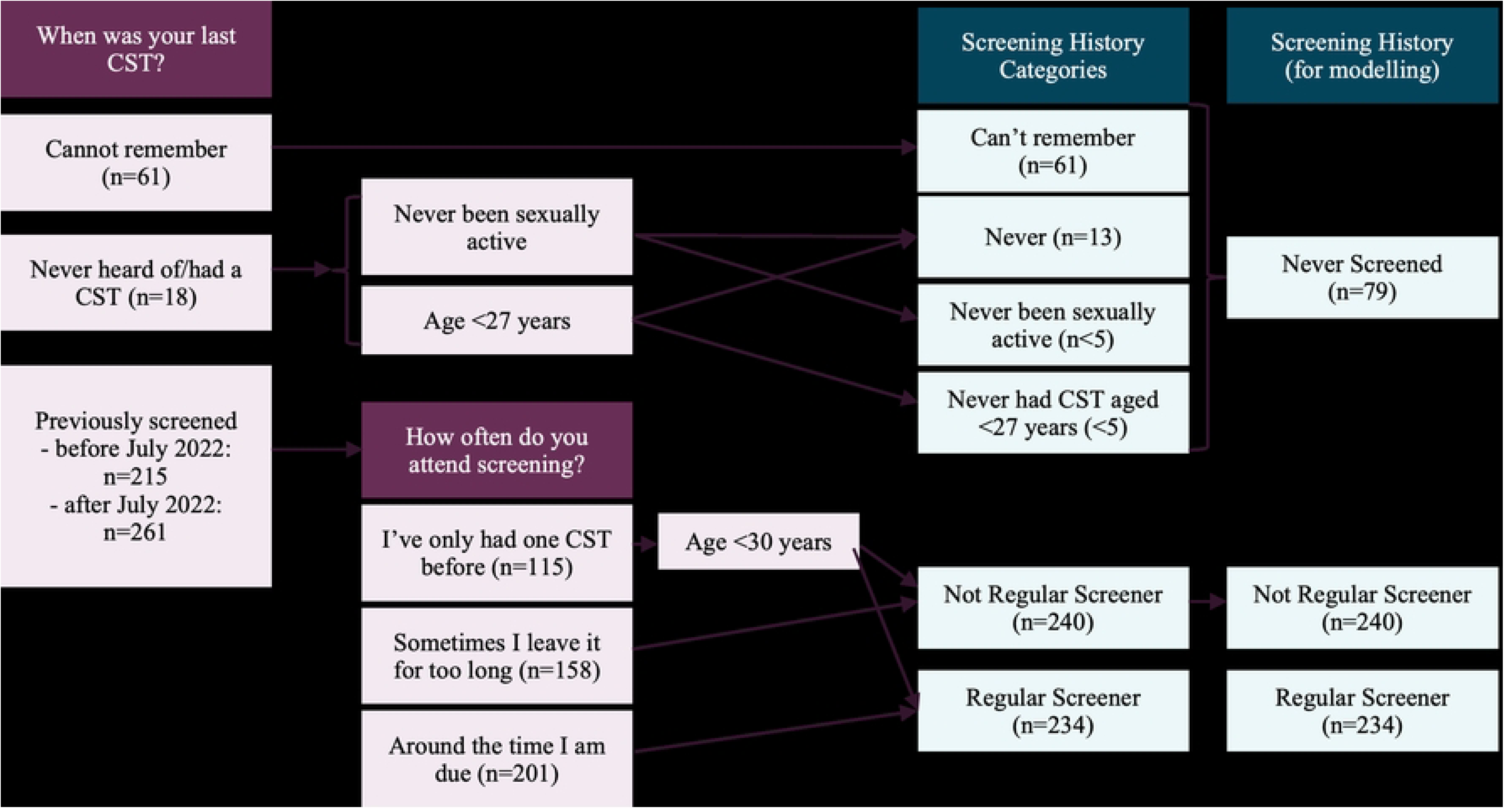
Process for categorisation of Screening History.

**Fig 2.**
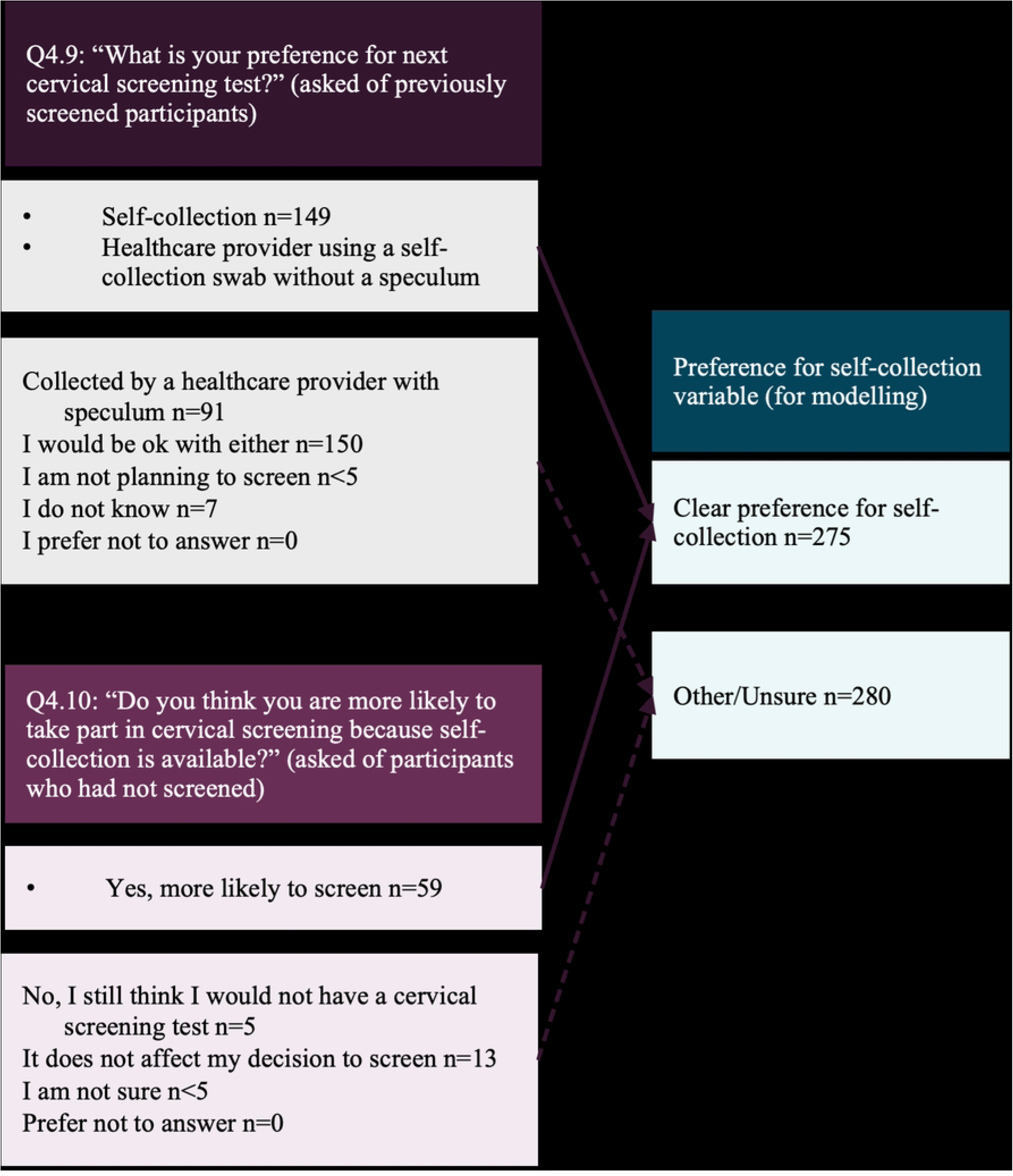
Process for categorisation of preferring self-collection.

Summary data are provided as number of observations (N) and percents (%). To examine the relationship of demographic and screening-related variables to future preference for self-collection (outcome variable), binomial logistic regression was performed. Model assumptions were checked, and where multicollinearity was a concern, one variable was selected *a priori* based on investigator judgment. Socioeconomic status was excluded from the model due to its limitations as a predictor among Aboriginal and/or Torres Strait Islander participants, particularly in urban areas (25), and collinearity with remoteness - both being postcode-derived. Unadjusted odds ratios (OR) were first calculated for each sociodemographic variable (age, sexuality, education, remoteness) and preference for self-collection, followed by adjusted odds ratios (adjOR) controlling for included predictors. To assess whether screening history mediates the relationship between sociodemographic factors and screening preference, we used a stepwise modelling approach comparing models excluding and including screening history. Mediation was evaluated by calculating the percentage change in odds ratios, with a 10% reduction considered indicative of mediation. Stepwise modelling indicated that screening history did not substantially mediate the relationship between sociodemographic factors and preference for self-collection (see Supporting Information – Modelling Output). The analysis is further described in Supporting Information – Analysis Plan. Data was analysed in STATA v18 (StataCorp, Texas, USA).

## Results

### Participants

A total of 555 screen eligible Aboriginal and Torres Strait Islander women and people with a cervix were included in the analysis. Participants predominantly identified as Aboriginal (87%), women (99%) and heterosexual (90%; Table 1). Almost half of the participants were aged 25-34 years of age (n=238, 44%), had tertiary qualifications (n=253, 50%), and lived in a major city (n=223, 56%). The majority (almost 85%) of survey participants had screened previously, classified as regular (n=234, 42%) or not-regular (n=240, 43%) screeners. A smaller proportion of participants (n=79, 14%) had never screened, and two did not include screening history and were therefore excluded.

**Table 1.**
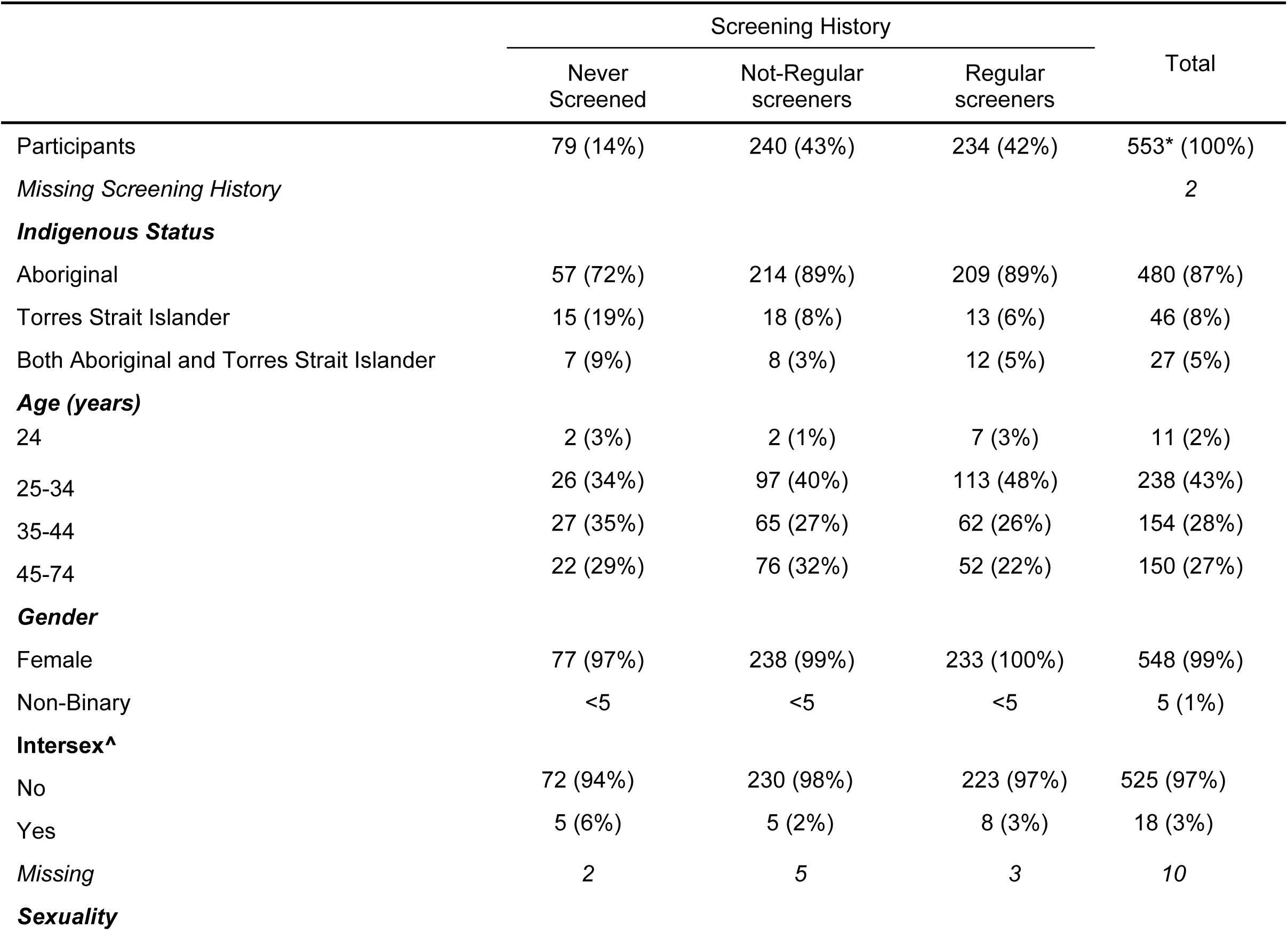
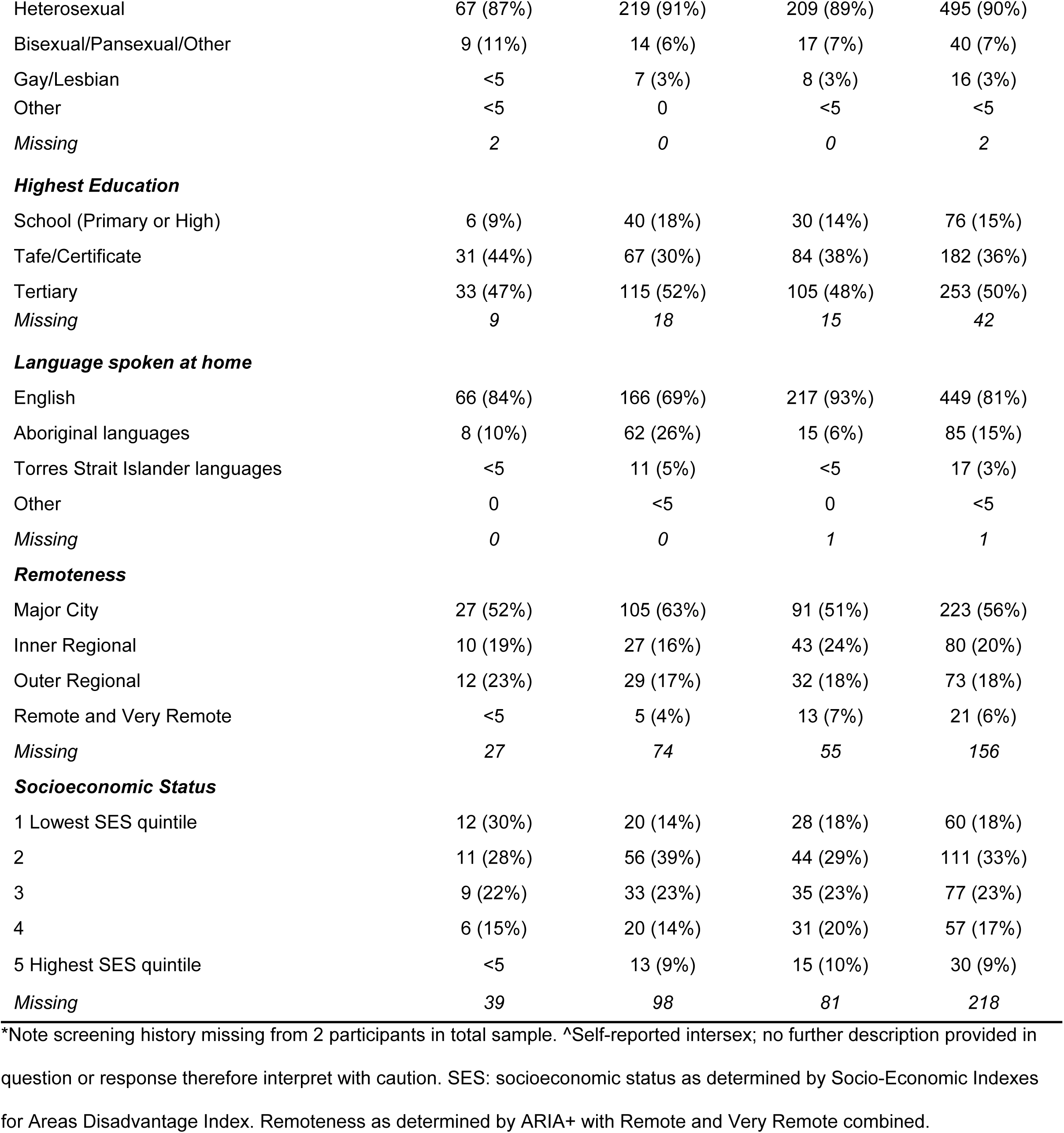
Participant characteristics by screening history.

|  | Screening History |  |  | Total |
| --- | --- | --- | --- | --- |
|  | Never Screened | Not-Regular screeners | Regular screeners |  |
| Participants | 79 (14%) | 240 (43%) | 234 (42%) | 553* (100%) |
| *Missing Screening History* |  |  |  | 2 |
| **Indigenous Status** |  |  |  |  |
| Aboriginal | 57 (72%) | 214 (89%) | 209 (89%) | 480 (87%) |
| Torres Strait Islander | 15 (19%) | 18 (8%) | 13 (6%) | 46 (8%) |
| Both Aboriginal and Torres Strait Islander | 7 (9%) | 8 (3%) | 12 (5%) | 27 (5%) |
| **Age (years)** |  |  |  |  |
| 24 | 2 (3%) | 2 (1%) | 7 (3%) | 11 (2%) |
| 25-34 | 26 (34%) | 97 (40%) | 113 (48%) | 238 (43%) |
| 35-44 | 27 (35%) | 65 (27%) | 62 (26%) | 154 (28%) |
| 45-74 | 22 (29%) | 76 (32%) | 52 (22%) | 150 (27%) |
| **Gender** |  |  |  |  |
| Female | 77 (97%) | 238 (99%) | 233 (100%) | 548 (99%) |
| Non-Binary | <5 | <5 | <5 | 5 (1%) |
| **Intersex^** |  |  |  |  |
| No | 72 (94%) | 230 (98%) | 223 (97%) | 525 (97%) |
| Yes | 5 (6%) | 5 (2%) | 8 (3%) | 18 (3%) |
| *Missing* | 2 | 5 | 3 | 10 |
| **Sexuality** |  |  |  |  |
| Heterosexual | 67 (87%) | 219 (91%) | 209 (89%) | 495 (90%) |
| Bisexual/Pansexual/Other | 9 (11%) | 14 (6%) | 17 (7%) | 40 (7%) |
| Gay/Lesbian | <5 | 7 (3%) | 8 (3%) | 16 (3%) |
| Other | <5 | 0 | <5 | <5 |
| *Missing* | 2 | 0 | 0 | 2 |
| ***Highest Education*** |  |  |  |  |
| School (Primary or High) | 6 (9%) | 40 (18%) | 30 (14%) | 76 (15%) |
| Tafe/Certificate | 31 (44%) | 67 (30%) | 84 (38%) | 182 (36%) |
| Tertiary | 33 (47%) | 115 (52%) | 105 (48%) | 253 (50%) |
| *Missing* | 9 | 18 | 15 | 42 |
| ***Language spoken at home*** |  |  |  |  |
| English | 66 (84%) | 166 (69%) | 217 (93%) | 449 (81%) |
| Aboriginal languages | 8 (10%) | 62 (26%) | 15 (6%) | 85 (15%) |
| Torres Strait Islander languages | <5 | 11 (5%) | <5 | 17 (3%) |
| Other | 0 | <5 | 0 | <5 |
| *Missing* | 0 | 0 | 1 | 1 |
| ***Remoteness*** |  |  |  |  |
| Major City | 27 (52%) | 105 (63%) | 91 (51%) | 223 (56%) |
| Inner Regional | 10 (19%) | 27 (16%) | 43 (24%) | 80 (20%) |
| Outer Regional | 12 (23%) | 29 (17%) | 32 (18%) | 73 (18%) |
| Remote and Very Remote | <5 | 5 (4%) | 13 (7%) | 21 (6%) |
| *Missing* | 27 | 74 | 55 | 156 |
| ***Socioeconomic Status*** |  |  |  |  |
| 1 Lowest SES quintile | 12 (30%) | 20 (14%) | 28 (18%) | 60 (18%) |
| 2 | 11 (28%) | 56 (39%) | 44 (29%) | 111 (33%) |
| 3 | 9 (22%) | 33 (23%) | 35 (23%) | 77 (23%) |
| 4 | 6 (15%) | 20 (14%) | 31 (20%) | 57 (17%) |
| 5 Highest SES quintile | <5 | 13 (9%) | 15 (10%) | 30 (9%) |
| *Missing* | 39 | 98 | 81 | 218 |

\*Note screening history missing from 2 participants in total sample. ^Self-reported intersex; no further description provided in question or response therefore interpret with caution. SES: socioeconomic status as determined by Socio-Economic Indexes for Areas Disadvantage Index. Remoteness as determined by ARIA+ with Remote and Very Remote combined.

### Awareness of self-collection

Most participants had heard of self-collection (n=435, 79%; Table 2), however, regular screeners were 4.0 times more likely to have heard of self-collection than those who had never screened (OR=4.0, 95%CI 2.3-7.0, p<0001). Previously screened participants, either regular or not-regular, most frequently cited healthcare providers as sources of knowledge, followed by conventional media and social media (Table 2). In contrast, never screened participants mostly reported traditional media (n=18, 23%) and social media (n=12, 15%) as sources rather than their healthcare provider (n=13, 16%).

**Table 2.** Awareness of and future preference for self-collection by screening history.

|  | Screening History |  |  | Total |
| --- | --- | --- | --- | --- |
|  | Never Screened | Not-Regular screeners | Regular screeners |  |
| Participants | 79 (14%) | 240 (43%) | 234 (42%) | 553* (100%) |
| ***Have you heard of self-collection?*** |  |  |  |  |
| Yes | 42 (53%) | 201 (84%) | 192 (82%) | 435 (79%) |
| No | 37 (47%) | 39 (16%) | 42 (18%) | 118 (21%) |
| Missing |  |  |  | 0 |
| ***If Yes, where did you hear about self-collection? (Select all that apply)\**** |  |  |  |  |
| Healthcare provider told me about it | 13 (16%) | 77 (32%) | 79 (34%) | 169 (31%) |
| Saw something in the media (television, radio, etc.) | 18 (23%) | 57 (24%) | 53 (23%) | 128 (23%) |
| Social media | 12 (15%) | 53 (22%) | 44 (19%) | 109 (20%) |
| Family or friend | 5 (6%) | 34 (14%) | 43 (18%) | 82 (15%) |
| Letter from the National Cancer Screening Register | 6 (8%) | 32 (13%) | 28 (12%) | 66 (12%) |
| A letter or phone call from my healthcare clinic | 0 | 40 (17%) | 22 (9%) | 62 (11%) |
| Website | 0 | 16 (7%) | 10 (4%) | 26 (5%) |
| Campaign from an organisation e.g. Cancer Council, the Government or ACON | <5 | <5 | 15 (6%) | 21 (4%) |
| Other | <5 | <5 | 15 (6%) | 21 (4%) |
| Unsure | 0 | <5 | <5 | <5 |
| ***How would you prefer to do your next CST?^*** |  |  |  |  |
| Self-collection | n.a. | 85 (35%) | 64 (27%) | 149 (27%) |
| HCP help without speculum | n.a. | 38 (16%) | 29 (12%) | 67 (12%) |
| HCP with speculum | n.a. | 33 (14%) | 58 (25%) | 91 (16%) |
| Either option | n.a. | 74 (31%) | 76 (32%) | 150 (27%) |
| Not planning to screen | n.a. | <5 | 0 | <5 |
| Unsure | n.a. | <5 | <5 | 7 (1%) |
| *Missing* |  | 2 | 4 | 6 |
| ***Are you more likely to screen because of self-collection?#*** |  |  |  |  |
| Yes, more likely to screen | 59 (75%) | n.a. | n.a. | 59 (75%) |
| Doesn't affect decision to screen | 13 (16%) | n.a. | n.a. | 13 (16%) |
| No, still would not screen / Unsure | 7 (9%) | n.a. | n.a. | 7 (9%) |
| *Missing* |  |  |  | 0 |

Note: \*Participants could choose multiple options where they heard of self-collection therefore percentages will not add up to 100%. ^This question was only asked of respondents who had previously participated in cervical screening. #This question was only asked of respondents who had not previously participated in cervical screening. CST: cervical screening test. n.a.survey participants in this category were not asked this question.

### Preference for self-collection

When asked how they would prefer to do their next cervical screening test, 27% of regular screeners (n=64) and 35% of not-regular screeners (n=85) preferred self-collection. An additional 12% (n=29) and 16% (n=38), respectively, preferred for a healthcare provider to assist them to collect a self-collected sample (i.e. without a speculum). Regular screeners were more likely to prefer having a clinician-collected sample (with a speculum, n=58, 25%) at their next screen compared to not-regular screeners (n=33, 14%). Approximately a third said they would be happy with either option (Table 2). In contrast, participants who had never screened indicated that they were more likely to take part in cervical screening because self-collection was available (n=59, 75%; Table 2).

Half of survey participants indicated a clear preference for self-collection, either collecting themselves or assisted by a healthcare provider without a speculum (n=275) and the other half indicated they would screen via clinician-collection, either option or were unsure (n=280, Table 3).

**Table 3.**
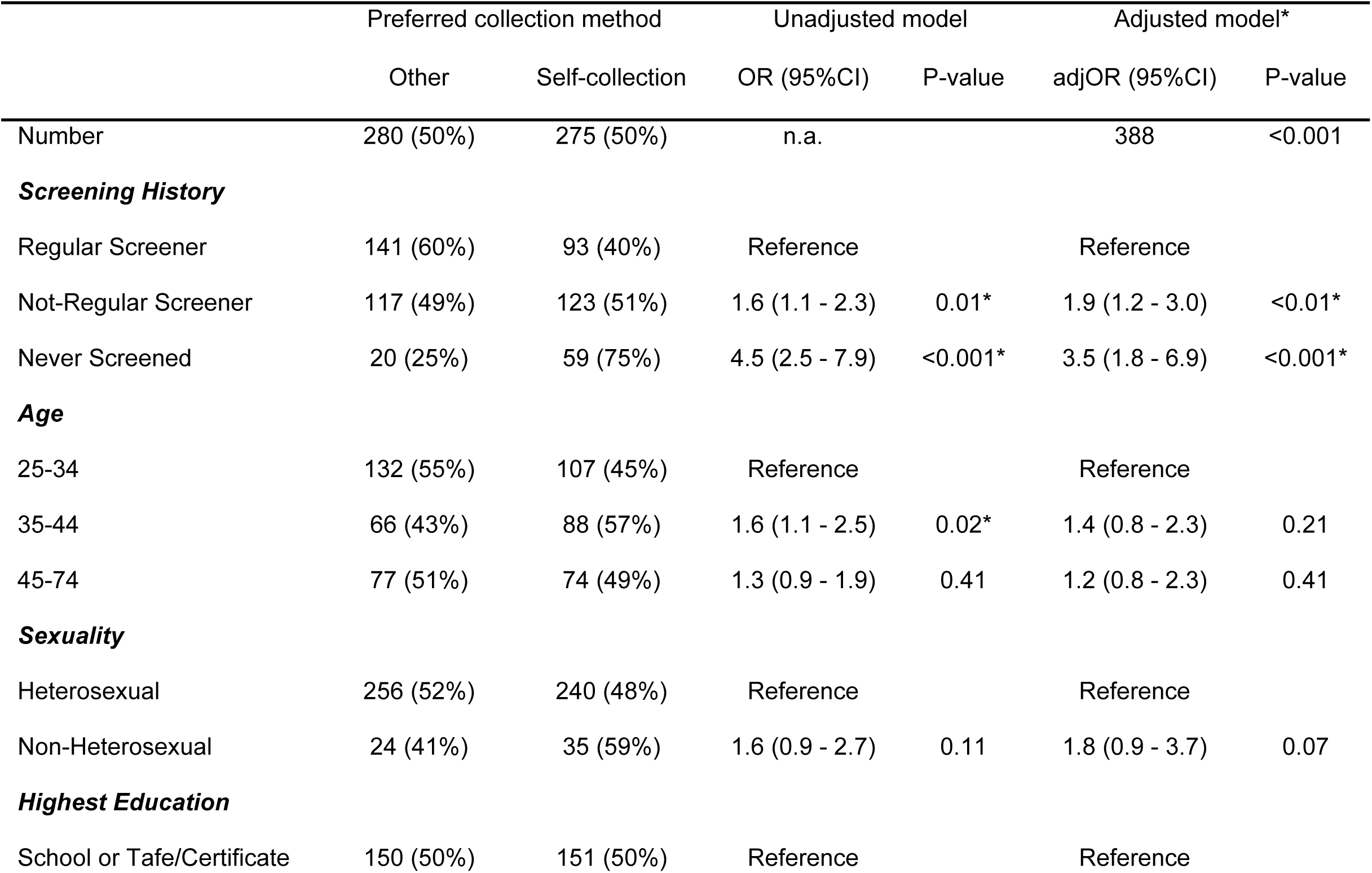
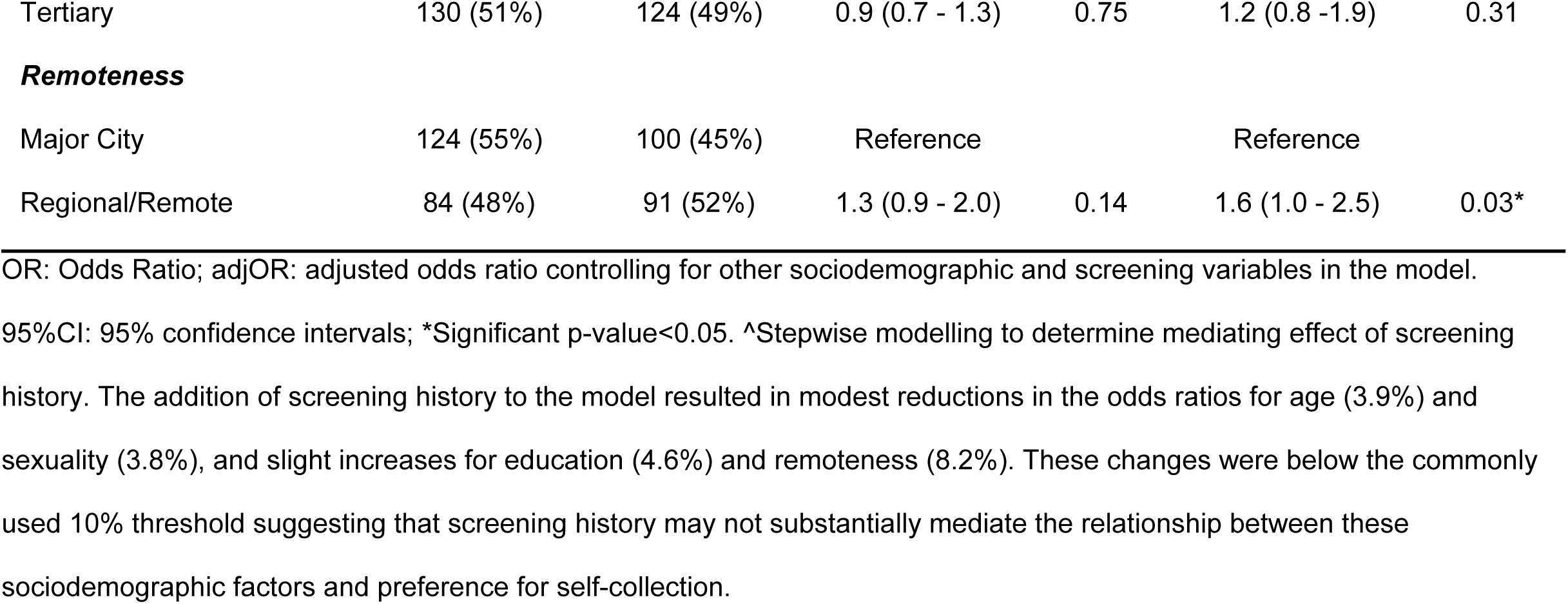
Factors associated with a preference for self-collection.

|  | Preferred collection method |  | Unadjusted model |  | Adjusted model* |  |
| --- | --- | --- | --- | --- | --- | --- |
|  | Other | Self-collection | OR (95%CI) | P-value | adjOR (95%CI) | P-value |
| Number | 280 (50%) | 275 (50%) | n.a. |  | 388 | <0.001 |
| **Screening History** |  |  |  |  |  |  |
| Regular Screener | 141 (60%) | 93 (40%) | Reference |  | Reference |  |
| Not-Regular Screener | 117 (49%) | 123 (51%) | 1.6 (1.1 - 2.3) | 0.01* | 1.9 (1.2 - 3.0) | <0.01* |
| Never Screened | 20 (25%) | 59 (75%) | 4.5 (2.5 - 7.9) | <0.001* | 3.5 (1.8 - 6.9) | <0.001* |
| **Age** |  |  |  |  |  |  |
| 25-34 | 132 (55%) | 107 (45%) | Reference |  | Reference |  |
| 35-44 | 66 (43%) | 88 (57%) | 1.6 (1.1 - 2.5) | 0.02* | 1.4 (0.8 - 2.3) | 0.21 |
| 45-74 | 77 (51%) | 74 (49%) | 1.3 (0.9 - 1.9) | 0.41 | 1.2 (0.8 - 2.3) | 0.41 |
| **Sexuality** |  |  |  |  |  |  |
| Heterosexual | 256 (52%) | 240 (48%) | Reference |  | Reference |  |
| Non-Heterosexual | 24 (41%) | 35 (59%) | 1.6 (0.9 - 2.7) | 0.11 | 1.8 (0.9 - 3.7) | 0.07 |
| **Highest Education** |  |  |  |  |  |  |
| School or Tafe/Certificate | 150 (50%) | 151 (50%) | Reference |  | Reference |  |
| Tertiary | 130 (51%) | 124 (49%) | 0.9 (0.7 - 1.3) | 0.75 | 1.2 (0.8 -1.9) | 0.31 |
| **Remoteness** |  |  |  |  |  |  |
| Major City | 124 (55%) | 100 (45%) | Reference |  | Reference |  |
| Regional/Remote | 84 (48%) | 91 (52%) | 1.3 (0.9 - 2.0) | 0.14 | 1.6 (1.0 - 2.5) | 0.03* |

OR: Odds Ratio; adjOR: adjusted odds ratio controlling for other sociodemographic and screening variables in the model.
95%CI: 95% confidence intervals; \*Significant p-value<0.05. ^Stepwise modelling to determine mediating effect of screening history. The addition of screening history to the model resulted in modest reductions in the odds ratios for age (3.9%) and sexuality (3.8%), and slight increases for education (4.6%) and remoteness (8.2%). These changes were below the commonly used 10% threshold suggesting that screening history may not substantially mediate the relationship between these sociodemographic factors and preference for self-collection.

Previous screening history was significantly associated with a preference for self-collection (Table 3). Compared to regular screeners, not-regular screeners were more likely to prefer self-collection (51% vs 40%; OR=1.6; 95%CI=1.1-2.3, p=0.01). After adjusting for other demographic variables, not-regular screeners were 1.9 times more likely to have a clear preference for self-collection (95%CI=1.2–3.0, p<0.01). This preference for self-collection was stronger among never-screened participants (75% vs 40%; OR=4.5; 95%CI= 2.5-7.9, p<0.001). After adjustment, never screened participants were 3.5 times more likely to have a clear preference for self-collection compared to regular screeners (95%CI=1.8-6.9, p<0.001).

There was a trend towards a preference among non-heterosexual participants, however confidence intervals were large due to small numbers meaning this was non-significant (adjOR=1.8, 95%CI= 0.9–3.7, p=0.07). Increasing age and education were not significantly associated with a preference with self-collection in our sample. Remoteness alone was not significantly associated with a preference with self-collection, however after controlling for other screening and demographic variables, participants in regional or remote areas were 1.6 times more likely (95%CI=1.0–2.5, p=0.03). Model diagnostics are available in Supporting Information.

## Discussion

To our knowledge, this is the first study to quantify awareness of and preference for self-collection among Aboriginal and Torres Strait Islander women and people with a cervix since the introduction of universal access to self-collection in Australia’s NCSP. In addition to reinforcing previous evidence among those who have screened, our findings provide new insights into preferences among never-screened participants, who were significantly more likely to prefer self-collection when aware of this option. While overall awareness of self-collection was high, this likely reflects a self-selected sample, most of whom had previously screened. In contrast, individuals who had never screened were significantly less aware of self-collection and less likely to have heard about it from a healthcare provider, despite being much more likely to prefer self-collection as an option. This highlights a key opportunity for providers to engage never-screened clients by discussing self-collection.

The frequent mention of both traditional and social media as information sources suggests that broad media campaigns can effectively reach those who don’t typically participate in cervical screening. The frequency of nominating social media may have been influenced by almost half of our sample being aged 25-34 years, and possibly more engaged with social media. Since our survey, a national media campaign, focused on both participants and providers, has been in market promoting awareness about cervical screening, and in particular the option to choose self-collection. (26) This campaign has included a targeted campaigns aimed at Aboriginal and Torres Strait Islander people. Evidence on the effectiveness of media campaigns in increasing cervical screening participation remains limited, particularly for Aboriginal and Torres Strait Islander women and other people with a cervix. Robust evaluation is essential to determine whether such initiatives genuinely support informed participation rather than simply increase awareness. While mass media campaigns have shown modest impacts in general populations, their reach and cultural appropriateness for Indigenous communities are untested. Emerging evidence suggests that social media interventions can enhance awareness and influence screening intentions, particularly when messages address known barriers. (27, 28), However, their effectiveness among never-screened Aboriginal and Torres Strait Islander populations remains unknown. This gap highlights the need for multi-channel, community-led strategies and rigorous evaluation frameworks that measure not only participation but also cultural safety, trust, and long-term engagement.

These findings also provide important context for understanding potential behaviour change. When asked about future screening preferences, 50% of survey participants had a clear preference for self-collection and 50% indicated that they would either prefer clinician-collection or had no preference (including some who were unsure). This shows a higher preference for self-collection among Aboriginal and Torres Strait Islander screen eligible participants surveyed than is seen in national screening data at the time of the survey, which was around 30% at the time (although these uptake data would represent a combination of preference and access to choice). (29) Since then, self-collection has increased to represent representing almost 46% of all HPV screening tests in Quarter 2-2025 - a substantial rise that suggests growing acceptance of self-collection nationally, aligning with preferences observed in our sample. Under-and never screened Aboriginal and Torres Strait Islander women and people with a cervix surveyed were significantly more likely to prefer self-collection and that self-collection would make them more likely to participate in cervical screening. This is consistent with previous literature showing self-collection is preferred and seen as a potential solution to some existing screening barriers among under- and never-screened Aboriginal and Torres Strait Islander women and people with a cervix (14, 16, 30), and consistent with national uptake of self-collection among under- and never-screened participants (54.4% and 60.2% respectively). (29) There is limited evidence on how best to engage under- and never-screened Aboriginal and Torres Strait Islander peoples in cervical screening. Current evidence, although limited, highlights the importance of implementing multiple, Indigenous-led strategies to improve engagement in cervical screening. Effective approaches include flexible models of care, culturally safe services, and co-designed education to increase health literacy and awareness of self-collection. (5)

Study participants living in regional or remote areas of Australia in our sample were more likely to express a preference for self-collection after controlling for other factors. This finding is consistent with national self-collection uptake data, with almost 67% of samples from screening participants in very remote areas being self-collected. (29) Access to self-collection in these areas is important as cervical screening participation rates are historically lower among those living more remote or those who are typically more socioeconomically disadvantaged. (3) Aboriginal and Torres Strait Islander women and people with a cervix in these areas face significant barriers, including workforce shortages and limited access to any recommended follow-up tests or treatment. (7, 31) Those who do screen often experience delays in receiving results and treatment due to geographical distances. Consequently, they are more likely to be diagnosed with advanced cancers and less likely to receive timely treatment compared to urban residents. (4) Self-collection supports the delivery of flexible models of care, such as telehealth, point-of-care testing, outreach and home-based screening, overcoming many previously identified practical barriers identified to attending or accessing screening, such as costs, transport, childcare or work commitments. (5, 15, 16, 32–35).

In our sample, other sociodemographic factors—such as age, sexuality, and education—were not significantly associated with a preference for self-collection. Instead, findings highlight that avoiding trauma, discomfort, and embarrassment is a shared concern, reinforcing the need to offer all eligible participants a choice in how they screen. reinforcing the importance of offering all eligible participants a choice in how they screen. This is particularly critical for those who are under- or never-screened, as avoidance is often driven by fear, embarrassment, or reluctance to be touched by a healthcare provider. (14, 16, 17, 30) While most participants identified as heterosexual females, there were indications of a stronger preference for self-collection among those identifying as non-heterosexual, highlighting the need for further research into the preferences of sexually and gender diverse Aboriginal and Torres Strait Islander people with a cervix. A preference for self-collection was also observed among participants aged 35–44 in the unadjusted model. Although specific data for Aboriginal and Torres Strait Islander groups are not available, national trends show increasing uptake of self-collection with age—highest among those aged 70–74 (53.5%) and lowest among 30–34-year-olds (42.7%) in Q2 2025. (29) These patterns suggest that age influences screening choices, emphasising the need for tailored communication strategies.

Age is also an important consideration for informed decision-making, as younger participants have higher rates of HPV (not 16/18), which often necessitates follow-up clinician-collected samples. (3, 36) Current guidelines recommend that informed choice includes discussion of the likelihood of follow-up testing (36), as acceptability of self-collection tends to decrease when additional testing is required. (7)

Finally, it is important to note that this survey did not explore the impacts of colonisation, racism, or access to culturally safe care—factors that are known to affect healthcare access. Understanding the facilitators of self-collection in the context of ongoing colonisation is essential for equitable cervical cancer elimination. While universal access will likely go some way to increase equity, these are likely to persist if self-collection is implemented in isolation and without systemic reforms that support culturally safe, strengths-based community-driven approaches to service provision.

## Strengths and Limitations

Our sample was typically women who had previously screened, were young, tertiary level educated and living in a major city. While this was accounted for within the adjusted model, this means that results may not be generalisable to the broader population, especially to never-screened Aboriginal and Torres Strait Islander peoples and those living in remote areas, where our sample was smaller. This study relied on online recruitment, which may have excluded individuals without reliable internet access and introduced sampling bias.

Additionally, while multiple measures were implemented to minimize fraudulent responses, online surveys remain vulnerable to such risks. Compared to the broader population, our sample over-represented participants aged 25–34 years and those living in major cities, while under-representing older age groups and people in very remote areas, likely reflecting the primary recruitment method.

Cervical screening history was self-reported and drawn from a convenience sample, which may not be representative and cannot be verified against clinical records, introducing potential recall bias. Finally, the study did not collect qualitative data on personal, cultural, or emotional factors influencing screening choices, or other factors beyond those included in the modelling. Future research should incorporate alternative recruitment strategies and explore qualitative insights to better understand barriers and preferences.

## Conclusions

There is a clear preference for self-collection among under-screened and never-screened Aboriginal and Torres Strait Islander women and people with a cervix. However, awareness of self-collection at the time of our survey was lower among those who had never been screened. This presents a valuable opportunity to improve cervical screening participation among Aboriginal and Torres Strait Islander women and people with a cervix by increasing awareness and accessibility of self-collection.

## Data Availability

In accordance with principles of Indigenous Data Sovereignty, the research team remains the custodian of the data, and Aboriginal and Torres Strait Islander participants retain ownership. The data collected in this study includes individual survey responses which contain potentially identifying and sensitive information. Participants did not consent to public release of their data, and access is subject to ethical restrictions imposed by the Human Research Ethics Committees (HRECs) listed in the Ethics Approval section. Requests for access to the minimal de identified dataset reviewed in consultation with Thiitu Tharrmay and may be directed to YardhuraWalani, Australian National University ( phone: +61261255111), or the Australian Institute of Aboriginal and Torres Strait Islander Studies (AIATSIS phone: +61262461111). Powered

## Acknowledgments

We wish to thank those who participated in the study. We acknowledge the assistance and guidance of the Screen Your Way Investigators, Aboriginal and Torres Strait Islander Caucus as well as Aboriginal and Torres Strait Islander reference group Thiitu Tharrmay. Membership of the Screen Your Way Investigator Team includes Chief Investigators: Associate Professor Julia Brotherton, Professor Gail Garvey, Dr Tamara Butler, Associate Professor Mark Wenitong, Associate Professor Megan Smith, Associate Professor Claire Nightingale, Professor Marion Saville, Professor Rebecca Guy, and Professor Joan Cunningham; and Associate Investigators: Claudette “Sissy” Tyson, Sonya Egert, Kristine Falzon, Professor Bev Lawton, Professor Karen Canfell, Associate Professor Natalie Taylor and Dr Hamish McManus. We also thank Josephine Mondino, Cancer Council Australia, for providing support and advice for the Meta recruitment campaign. Ownership of Aboriginal and Torres Strait Islander knowledges and cultural heritage is retained by the informants.

## Abbreviations

CST: cervical screening test
HPV: Human Papillomavirus
NCSP: National Cervical Screening Program
OR: Odds Ratio
adjOR: Adjusted Odds Ratio
95%CI: 95% confidence intervals

## Authors contributions

All authors contributed to the conception, design, and development of this study. Each author participated in the survey design, coding and data analysis, drafting the manuscript, revising it critically for important intellectual content, and approving the final version to be published. All authors agree to be accountable for all aspects of the work.

## Data sharing statement

All authors had full access to all the data (including statistical reports and tables) related to the study. The data for this study will not be shared broadly but would require ethical approval to do so. Responses to survey questions relating to items relevant to these analyses are provided in the Supplemental Information, with counts suppressed where relevant to maintain participant privacy.

## Funding

This research has been funded through two Australian National Health and Medical Research Council (NHMRC) Targeted Call for Research competitive funding grants (GNT201490, GNT201578). LJW is supported by a NHMRC Investigator Grant (2009380) CN is supported by a Mid-Career Research Fellowship (MCRF21039) from the Victorian Government acting through the Victorian Cancer Agency. CJJ is supported by a University of Sydney PhD scholarship. The funding organisations have no role in the collection, management, analysis, and interpretation of data; writing of the protocol or subsequent manuscripts; and the decision to submit the protocol for publication. The views expressed in this publication are those of the authors and do not necessarily reflect the views of the funders.

## Competing interests

Nil

## Patient and public involvement

Patients and/or the public were not involved in the conduct, or reporting, or dissemination plans of this research. Pilot testing of the participant information and consent forms and survey was completed by a consumer advisory panel.

External Aboriginal and Torres Strait Islander Governance group Thiitu Tharrmay provided advice on recruitment and sampling.

## Patient consent for publication

Provided at time of patient consent.

## Provenance and peer review

Not commissioned; externally peer reviewed through grant and ethics processes.

## Notes

### Competing Interest Statement

The authors have declared no competing interest.

### Funding Statement

Yes

### Author Declarations

Ethics approval was obtained from the Australian Institute of Aboriginal and Torres Strait Islander Studies Research Ethics Committee (REC-0209) and ACON (RERC 202404), and was ratified by The Australian National University (H/2023/1434) and the University of Melbourne (2023-28454-47328-1) Human Research Ethics Committees. Written informed consent was obtained from all study participants. Before beginning the survey, participants were shown an embedded consent form within Qualtrics. Informed consent was obtained by asking participants the question: “Would you like to take part in this survey?” Only those who selected “I agree to take part in this survey” were able to proceed, thereby indicating their consent to participate.

